# State- and County-Level COVID-19 Public Health Orders in California: Constructing a Dataset and Describing Their Timing, Content, and Stricture

**DOI:** 10.1101/2020.11.08.20224915

**Authors:** Jeremy D. Goldhaber-Fiebert, Alexander F. Holsinger, Erin Holsinger, Elizabeth Long, on behalf of the SC-COSMO Modeling Consortium

## Abstract

Without vaccines, non-pharmaceutical interventions have been the most widely used approach to controlling the spread of COVID-19 epidemics. Various jurisdictions have implemented public health orders as a means of reducing effective contacts and controlling their local epidemics. Multiple studies have examined the effectiveness of various orders (e.g. use of face masks) for epidemic control. However, orders occur at different timings across jurisdictions and some orders on the same topic are stricter than others. We constructed a county-level longitudinal data set of more than 2,400 public health orders issues by California and its 58 counties pertaining to its 40 million residents. First, we describe methods used to construct the dataset that enables the characterization of the evolution over time of California state- and county-level public health orders dealing with COVID-19 from January 1, 2020 through June 30, 2021. Public health orders are both an interesting and important outcome in their own right and also a key input into analyses looking at how such orders may impact COVID-19 epidemics. To construct the dataset, we developed and executed a search strategy to identify COVID-19 public health orders over this time period for all relevant jurisdictions. We characterized each identified public health order in terms of the timing of when it was announced, went into effect and (potentially) expired. We also adapted an existing schema to describe the topic(s) each public health order dealt with and the level of stricture each imposed, applying it to all identified orders. Finally, as an initial assessment, we examined the patterns of public health orders within and across counties, focusing on the timing of orders, the rate of increase and decrease in stricture, and on variation and convergence of orders within regions.

## Public Availability

We are making the dataset publicly available as part of our commitment to open data and combating the COVID-19 pandemic (http://sc-cosmo.org). We intend to update the dataset at least monthly. We are hopeful that others will use our methodological approach to characterize county-level or even city-level public health orders over time across additional states and would be happy to discuss this further via email. A description of the contents and linkages of released files are provided in Appendix Table 1.

### Public Health Order Searches

We developed and executed a search strategy to identify COVID-19 public health orders over the analytic time period applied to all California counties and the state itself. We identified website(s) for each county on which public health orders were released and supplemented this with web searches for press releases and media that combined county name, terms relating to COVID-19, and terms relating to public health orders and the topics they dealt with [1-59] (see also Appendix Tables 2 and 3). Search terms used in Google included combinations of each county’s name and any of the following: declares emergency; bans gatherings; closes March; reopens April; reopens May; reopens June; Stage 2; Stage 3; recloses July; face coverings.

We recorded URLs and the date that each document was accessed, also archiving documents whenever possible. As some counties maintain only their most recent public health order on their websites (e.g., Solano County), we used the Wayback Machine to examine the orders contained on each such website at all points in time at which orders were added or changed [60]. In addition to gathering county public health orders, we also searched for and compiled the timing and topics of state-level public health guidance that applied to some or all of counties (e.g., all counties in the state, counties on a watch list, etc.) as this sometimes, explicitly or implicitly, substituted for or superseded county orders. Finally, when we encountered them, we separately noted instances where other county ordinances and orders may have related to COVID-19 but were not explicitly public health orders; we used this last category of information for narrative purposes only, excluding it from our dataset and analyses described below as we could not be sure that our search strategy would systematically identifying them in an equivalent way across time and counties.

### Characteristic Extraction

All public health orders and guidance had the following characteristics: 1) county(s) to which they pertained; 2) date of announcement; 3) date of effect; and 4) date of expiry^1^ (which could be either an explicit date or ongoing).

### Classification of Public Health Order Topics

We began the schema for order classification using topics found in the COVID-19 US State Policy Database [61]. To these, we added 5 topics to reflect more thoroughly the actions found in the California county public health orders: moratorium on gatherings; quarantine for residents; and lab reporting & contact tracing; characterization of social bubbling; and details of private events. With the introduction of vaccines to California in December 2020, we split appropriate measures into those for vaccinated and unvaccinated individuals. The complete list of topics and their general definition are provided in Appendix Table 4. Based on full textual review by a study investigator, we classified each identified public health order as to whether it pertained to each of the 16 main topics (e.g., face masks, gathering size, etc.). In the event of ambiguity of whether an order dealt with a given topic, a second study investigator reviewed the text and discussion to reach consensus was used to reach a final determination. A composite category of current level of county closure was added to the list which combined initial closures, reopenings, and reclosures (see below for further detail).

### Classification of Stricture on Each Public Health Order Topic

The primary focus of the classification of stricture was to gauge how public health orders relating to specific topics became more or less strict within each county over time. For example, instead of attempting to rank whether closure of indoor dining in a given county was more or less strict than closure of nail salons in another county, we focused on how the set of business types that were closed in a given county widened or narrowed over time. For certain topic areas (e.g., state of emergency), stricture was binary – whether an order about it existed at a given point in time or not. For others, stricture had ordinality (e.g., the increasing number of people allowed at gatherings or events). In addition to applying a within-county numeric scale, specific free text information was extracted to capture the details of how the order’s stricture on a given topic was determined. Lists of scales and their definitions are provided in Appendix Table 5. Based upon the scales we used, a study investigator classified the strictness of each order for each category it pertained to. Again, in the event of ambiguity, a second study investigator reviewed and discussed the particular example until consensus was reached. Since public health orders dealt with closures, reopenings, and reclosures, we constructed a combined variable on current strictness of closure/openness (which essentially added closure, reopening, and reclosure with a floor of 0 for completely open to account for differences in the ordinal scales we used). The main determinant of this variable’s value became the state-level tiered system upon the system’s rollout on August 31^st^ with the exception that counties could opt to remain more strictly closed than the state system required [62].

The use of expiration dates issued with each order varies by order topic. Topics related to opening and closing, for example, physical distancing, stay at home, reopening, reclosing, and social bubbles tended to have expiration dates issued with orders. In many cases the orders were superseded by new orders prior to the expiration dates, but expiration dates are honored for these orders if they were not superseded. These expirations explain the numerous changes to closure and stay at home levels in January, despite the low number of orders for the month. Lab and contact tracing, mask restrictions, and quarantine restrictions tended to not have expiration dates published with the orders and are therefore only modified in the data if they have a new order that specifically increases or decreases the stricture of the previous order.

### Descriptive Analysis of Public Health Orders and Their Topics and Stricture

We identified and characterized 2,457 state- and county-level public health orders related to COVID-19 that were issued and became effective between January 1, 2020 and June 30, 2021. Of those, 2,268 dealt with the key topics of physical distancing, stay at home, gatherings for unvaccinated individuals, gatherings for fully vaccinated individuals, private events for unvaccinated individuals, private events for fully vaccinated individuals, quarantine instructions for unvaccinated residents, quarantine instructions for fully vaccinated residents, quarantine instructions for unvaccinated visitors, quarantine instructions for fully vaccinated visitors, reopening, reclosing, lab and contact tracing, mask restrictions, social bubbles for unvaccinated households, and social bubbles for fully vaccinated households^2,3,4^.

#### Counts of orders

For the date ranges reviewed, we determined the number of COVID-19 public health orders promulgated. The months with the largest numbers of orders corresponded to rises in detected case rates.

**Table 1:** Number of orders by month

| Month | Number of distinct orders |
| --- | --- |
| January | 3 |
| February | 5 |
| March | 273 |
| April | 154 |
| May | 239 |
| June | 221 |
| July | 223 |
| August | 98 |
| September | 74 |
| October | 120 |
| November | 207 |
| December | 118 |
| January | 32 |
| February | 41 |
| March | 93 |
| April | 132 |
| May | 146 |
| June | 89 |

#### Counts of topics covered by orders

For the date ranges reviewed, we determined the number of order-topics (i.e., each time any COVID-19 public health order dealt with a given topic). Like counts of orders, we treated the topics covered by state guidance and orders in two ways: including them as a county orders when relevant or excluding them. We also calculated the percent of each specific order type as a function of total orders. Since some orders dealt with multiple topics in a single order, the percentages sum to greater than one. A large fraction of orders dealt with which venues would be open or closed and/or the levels of their closure/openness.

**Table 2:** Number of orders by topic and percent of orders by topic

| Topic | Number of orders | Percent of orders related to topic |
| --- | --- | --- |
| Gatherings for unvaccinated individuals | 218 | 9.6% |
| Gatherings for fully vaccinated individuals | 158 | 7.0% |
| Private events for unvaccinated individuals | 158 | 7.0% |
| Private events for fully vaccinated individuals | 158 | 7.0% |
| Physical distancing | 239 | 10.5% |
| Stay at home | 353 | 15.6% |
| Quarantine for unvaccinated residents | 241 | 10.6% |
| Quarantine for fully vaccinated residents | 47 | 2.1% |
| Quarantine for unvaccinated visitors | 47 | 2.1% |
| Quarantine for fully vaccinated visitors | 29 | 1.3% |
| Reopening | 926 | 40.8% |
| Reclosing | 201 | 8.9% |
| Lab and contact tracing | 94 | 4.1% |
| Mask restrictions for unvaccinated individuals | 470 | 20.7% |
| Mask restrictions for vaccinated individuals | 200 | 8.8% |
| Social bubbles for unvaccinated households | 545 | 24.0% |
| Social bubbles for fully vaccinated households | 189 | 8.3% |

#### Counts of orders by county

For the date ranges reviewed, we determined the number of COVID-19 public health orders promulgated by each county and examined the distribution of these counts across counties. Since state guidance and orders could substitute for or supersede county orders, we generated county-level counts and other descriptive statistics in two ways, including relevant state orders as a county order when it applied and excluding state orders in counts if they did not apply.

**Table 3:** Number of orders by county by month

| County | Mar | Apr | May | Jun | Jul | Aug | Sep | Oct | Nov | Dec | Jan | Feb | Mar | Apr | May | Jun |
| --- | --- | --- | --- | --- | --- | --- | --- | --- | --- | --- | --- | --- | --- | --- | --- | --- |
| Alameda | 7 | 4 | 2 | 3 | 6 | 2 | 2 | 5 | 5 | 2 | 1 | 2 | 2 | 2 | 2 | 3 |
| Alpine | 6 | 1 | 2 | 5 | 1 | 1 | 0 | 1 | 4 | 1 | 0 | 1 | 2 | 3 | 2 | 1 |
| Amador | 7 | 2 | 7 | 3 | 1 | 4 | 2 | 1 | 3 | 2 | 2 | 0 | 1 | 2 | 3 | 1 |
| Butte | 3 | 1 | 4 | 3 | 2 | 1 | 2 | 2 | 4 | 3 | 0 | 1 | 2 | 2 | 2 | 1 |
| Calaveras | 3 | 1 | 4 | 3 | 1 | 2 | 1 | 2 | 5 | 2 | 1 | 0 | 1 | 2 | 2 | 1 |
| Colusa | 1 | 0 | 4 | 2 | 3 | 2 | 1 | 2 | 5 | 1 | 2 | 0 | 3 | 1 | 2 | 1 |
| Contra Costa | 7 | 2 | 4 | 4 | 8 | 1 | 4 | 3 | 6 | 3 | 1 | 1 | 2 | 3 | 2 | 1 |
| Del Norte | 6 | 2 | 3 | 6 | 3 | 1 | 0 | 1 | 4 | 1 | 1 | 1 | 0 | 1 | 3 | 1 |
| El Dorado | 4 | 2 | 3 | 3 | 1 | 1 | 1 | 1 | 5 | 3 | 0 | 0 | 1 | 4 | 2 | 1 |
| Fresno | 2 | 1 | 5 | 4 | 5 | 1 | 2 | 1 | 4 | 1 | 0 | 1 | 1 | 3 | 2 | 1 |
| Glenn | 4 | 0 | 2 | 5 | 4 | 2 | 1 | 2 | 4 | 1 | 0 | 0 | 1 | 2 | 2 | 1 |
| Humboldt | 3 | 1 | 3 | 4 | 1 | 2 | 1 | 3 | 3 | 2 | 1 | 2 | 2 | 3 | 2 | 2 |
| Imperial | 2 | 1 | 4 | 5 | 6 | 2 | 1 | 3 | 2 | 2 | 0 | 1 | 3 | 3 | 2 | 2 |
| Inyo | 4 | 3 | 6 | 3 | 2 | 2 | 1 | 2 | 2 | 2 | 1 | 0 | 0 | 2 | 4 | 1 |
| Kern | 3 | 3 | 2 | 4 | 3 | 1 | 1 | 2 | 4 | 1 | 0 | 0 | 1 | 2 | 2 | 1 |
| Kings | 4 | 1 | 3 | 4 | 6 | 1 | 1 | 2 | 4 | 3 | 0 | 0 | 1 | 2 | 2 | 1 |
| Lake | 9 | 7 | 5 | 4 | 2 | 1 | 0 | 2 | 3 | 1 | 1 | 0 | 1 | 3 | 2 | 1 |
| Lassen | 2 | 3 | 5 | 3 | 1 | 1 | 1 | 1 | 3 | 1 | 1 | 0 | 2 | 2 | 2 | 1 |
| Los Angeles | 9 | 3 | 6 | 6 | 8 | 1 | 2 | 7 | 4 | 3 | 2 | 3 | 2 | 5 | 2 | 3 |
| Madera | 3 | 2 | 2 | 5 | 5 | 1 | 1 | 1 | 2 | 2 | 0 | 0 | 1 | 2 | 3 | 1 |
| Marin | 8 | 2 | 6 | 5 | 5 | 2 | 1 | 3 | 3 | 3 | 0 | 1 | 1 | 3 | 2 | 3 |
| Mariposa | 4 | 3 | 3 | 3 | 1 | 3 | 2 | 1 | 4 | 3 | 1 | 0 | 1 | 2 | 3 | 1 |
| Mendocino | 5 | 4 | 4 | 3 | 5 | 4 | 2 | 3 | 4 | 3 | 1 | 0 | 3 | 5 | 3 | 4 |
| Merced | 3 | 5 | 2 | 3 | 7 | 2 | 1 | 2 | 3 | 1 | 0 | 1 | 1 | 3 | 3 | 1 |
| Modoc | 3 | 0 | 3 | 3 | 4 | 2 | 0 | 1 | 5 | 1 | 1 | 0 | 3 | 3 | 2 | 1 |
| Mono | 5 | 3 | 6 | 4 | 6 | 2 | 1 | 2 | 1 | 4 | 3 | 0 | 2 | 4 | 3 | 1 |
| Monterey | 2 | 3 | 4 | 4 | 4 | 1 | 1 | 1 | 1 | 3 | 0 | 0 | 1 | 3 | 3 | 2 |
| Napa | 5 | 2 | 3 | 7 | 5 | 1 | 0 | 1 | 3 | 2 | 0 | 0 | 2 | 2 | 2 | 2 |
| Nevada | 4 | 5 | 4 | 3 | 3 | 1 | 1 | 1 | 4 | 2 | 0 | 1 | 2 | 2 | 2 | 2 |
| Orange | 4 | 3 | 4 | 4 | 6 | 1 | 1 | 1 | 5 | 2 | 0 | 0 | 3 | 1 | 3 | 2 |
| Placer | 6 | 3 | 4 | 3 | 2 | 1 | 1 | 2 | 5 | 3 | 0 | 1 | 1 | 2 | 4 | 1 |
| Plumas | 1 | 1 | 5 | 4 | 1 | 1 | 0 | 2 | 5 | 2 | 1 | 1 | 1 | 2 | 2 | 1 |
| Riverside | 10 | 9 | 5 | 4 | 5 | 1 | 1 | 2 | 2 | 1 | 0 | 0 | 1 | 2 | 2 | 1 |
| Sacramento | 3 | 1 | 3 | 4 | 4 | 4 | 3 | 2 | 4 | 2 | 2 | 2 | 5 | 3 | 2 | 3 |
| San Benito | 4 | 2 | 3 | 3 | 4 | 1 | 1 | 2 | 4 | 1 | 0 | 0 | 1 | 2 | 2 | 2 |
| San Bernardino | 6 | 5 | 4 | 1 | 5 | 1 | 1 | 1 | 2 | 1 | 0 | 1 | 2 | 3 | 2 | 1 |
| San Diego | 7 | 5 | 4 | 4 | 7 | 3 | 1 | 1 | 3 | 3 | 1 | 3 | 1 | 2 | 3 | 3 |
| San Francisco | 10 | 3 | 5 | 5 | 6 | 2 | 5 | 6 | 4 | 7 | 2 | 2 | 2 | 2 | 3 | 2 |
| San Joaquin | 8 | 4 | 5 | 4 | 5 | 1 | 2 | 1 | 4 | 1 | 0 | 1 | 0 | 2 | 2 | 3 |
| San Luis Obispo | 7 | 5 | 4 | 3 | 4 | 1 | 1 | 3 | 4 | 1 | 0 | 1 | 2 | 2 | 2 | 2 |
| San Mateo | 10 | 6 | 8 | 5 | 2 | 3 | 1 | 5 | 3 | 1 | 1 | 1 | 1 | 2 | 4 | 1 |
| Santa Barbara | 3 | 5 | 4 | 5 | 7 | 3 | 4 | 3 | 3 | 4 | 3 | 2 | 5 | 3 | 3 | 3 |
| Santa Clara | 8 | 3 | 1 | 3 | 7 | 2 | 1 | 3 | 4 | 3 | 1 | 3 | 2 | 1 | 5 | 2 |
| Santa Cruz | 6 | 3 | 3 | 5 | 5 | 2 | 2 | 2 | 4 | 1 | 1 | 0 | 2 | 2 | 4 | 1 |
| Shasta | 1 | 0 | 4 | 3 | 1 | 1 | 1 | 3 | 3 | 2 | 0 | 2 | 0 | 2 | 2 | 1 |
| Sierra | 3 | 1 | 5 | 4 | 1 | 4 | 1 | 2 | 2 | 1 | 0 | 1 | 1 | 3 | 2 | 1 |
| Siskiyou | 3 | 3 | 3 | 3 | 1 | 2 | 0 | 2 | 5 | 1 | 0 | 0 | 2 | 3 | 2 | 1 |
| Solano | 5 | 2 | 4 | 5 | 5 | 1 | 2 | 1 | 4 | 2 | 0 | 0 | 1 | 1 | 2 | 2 |
| Sonoma | 7 | 5 | 6 | 5 | 4 | 2 | 1 | 1 | 1 | 2 | 0 | 0 | 1 | 2 | 3 | 1 |
| Stanislaus | 4 | 3 | 6 | 3 | 4 | 1 | 1 | 2 | 4 | 2 | 0 | 0 | 2 | 1 | 2 | 2 |
| Sutter | 2 | 1 | 3 | 2 | 4 | 1 | 1 | 2 | 4 | 3 | 0 | 0 | 1 | 2 | 3 | 1 |
| Tehama | 2 | 0 | 3 | 3 | 1 | 2 | 1 | 2 | 2 | 2 | 0 | 0 | 1 | 1 | 4 | 1 |
| Trinity | 6 | 1 | 7 | 3 | 1 | 1 | 0 | 2 | 5 | 1 | 0 | 2 | 1 | 1 | 3 | 1 |
| Tulare | 6 | 3 | 6 | 4 | 4 | 1 | 1 | 1 | 2 | 1 | 0 | 0 | 1 | 2 | 2 | 2 |
| Tuolumne | 4 | 3 | 5 | 4 | 2 | 2 | 2 | 1 | 4 | 2 | 0 | 0 | 2 | 1 | 2 | 1 |
| Ventura | 4 | 3 | 5 | 5 | 5 | 3 | 1 | 2 | 4 | 1 | 0 | 0 | 1 | 2 | 2 | 2 |
| Yolo | 3 | 3 | 6 | 4 | 7 | 1 | 2 | 2 | 3 | 3 | 0 | 2 | 4 | 2 | 3 | 1 |
| Yuba | 2 | 1 | 3 | 2 | 4 | 1 | 1 | 2 | 4 | 3 | 0 | 0 | 1 | 1 | 3 | 1 |

#### Daily rates of public health order/topic stricture by county

We employed graphical analysis to characterize the timing and relative degree of stricture of each county’s orders related to each of the 16 topics. Additionally, we employed graphical analysis to characterize the relative degree of stricture within California regions. One particularly interesting feature that varies across counties is the use of face masking orders and the use of closure orders in the late spring and summer. In many counties, closure order strictness increased prior to masking order strictness. However, once masking order strictness increased, some counties appear to have moved to re-open while others maintained or increased closure strictness. The use of different order types as substitutes for one another or as complements is an important area for further research.

## Discussion and Conclusions

Prior to vaccines, non-pharmaceutical interventions were the most widely used approach to controlling the spread of COVID-19 epidemics. Various jurisdictions implemented public health orders as a means of reducing effective contacts and controlling their local epidemics. While multiple studies have examined the effectiveness of various orders (e.g. use of face masks) for epidemic control [63-69], a complete picture of the magnitude and mechanisms of effectiveness of orders and regulation will likely require geographical and temporal variation in the contents of orders and their strictness. Current data sets tend to report on orders and regulations on a national and/or state level with current county-level efforts not necessarily standardized in their review and curation nor in their characterization of the topics and level of strictness contained in each order [70-72].

We constructed a county-level longitudinal data set of public health orders issues by California and its 58 counties pertaining to its 40 million residents. We observe both spatial and temporal patterns in the timing and strictness of orders on a variety of topics across counties and over time. We have made the dataset publicly available as part of our commitment to open data and combating the COVID-19 pandemic (http://sc-cosmo.org).

There are many potential uses of the data set we have constructed and released. It can be used in a variety of quasi-experimental designs to estimate the effects of such orders on epidemic health outcomes and on non-health outcomes (e.g., unemployment), potentially refining or extending prior studies in this area. It can be used to examine mechanisms like behavioral responses to such orders and compliance using either surveys such as those whose results are made available by Facebook or measured behavioral responses like those made available by Google’s Mobility Trends [73-74].

The approach we have employed for California is not exclusively applicable to California, and our hope is that other researchers in other jurisdictions may build up similar data sets and make them publicly available, leveraging the methods and approach we have taken. We are eager to assist in such efforts to the extent possible.

In conclusion, our data set provides an open source and useful contribution to analyses of important issues related to the COVID-19 pandemic. Its methods may be used more broadly to characterize policy responses. Illustrating its utility, a descriptive examination of public health orders at the county-level in California that accounts for the timing of the orders, the topics the orders address, and their level of strictness highlights intriguing patterns like the use of masking orders as a complement to other types of closures in some jurisdictions, presumably attempting to further reduce COVID-19 incidence, and as a substitute to other types of closure in other jurisdictions, presumably attempt to maintain COVID-19 at low incidence while mitigating the effects of health orders on the local economy. Further analyses of such phenomena are highly relevant and urgently needed as the pandemic continues.

## Supporting information

Archive containing all csv files described in Appendix Table 1 as well as information for other Appendix Tables as relevant

## Data Availability

We are making the complete dataset publicly available as part of our commitment to open data and combating the COVID-19 pandemic. We have updated the dataset approximately monthly as we continued our review and extraction of public health orders.

https://sc-cosmo.org

## Appendix

**Appendix Table 1:** Details of released data files

|  |  |
| --- | --- |
| PH_Orders_long | Contains the main data that is the result of scraping the web for all county public health orders and state public health directives. Based on review of the full text, the orders are each coded as to which category(s) (e.g., masking, closures) they pertain to and the ordinal relative level of stricture they represent. The file contains the following columns: county, dt_PHO, order_value, order_type. The variable county refers to one of 58 of California's counties (without the trailing string "County"). The variable order_type refers to the category of restriction referred to in a public health order. The variable order_value refers to the level of stricture for a particular order type in a particular county. The variable dt_PHO refers to the date that the county was at a particular level of stricture for a particular order type. This data set does not directly show when the county put out each of its order but changes in levels of order_value for order type on consecutive days show when there was an order(s) propagated. |
| PH_Orders_order_type | Contains slightly longer textual descriptions of the order types. It can be joined with PH_Orders_long using the order_type column. |
| PH_Orders_order_value | Contains the meanings of the scales for order values in within each order type. It can be joined with PH_Orders_order_type using the order_type column or with the PH_Orders_long using the order_type and order_value columns. |
| PH_Orders_region | Contains groupings of counties into regions based on CDPH groupings for "Economic Regions". Can be found at: calcat.covid19.ca.gov/cacovidmodels/ |
| PH_Orders_statewide | Contains information when state-wide public health orders came into effect. It essentially shows which order type the state-wide |
|  | order pertains to and the date on which it went into effect. It also documents the order value that was mandated. Note several important things. First, when the state produced tiers, the value for some counties could differ from others depending on the tiers they were in (hence in the order_value column for the statewide file we see a set of values that correspond to the state-mandated levels for each tier). Second, counties could opt to remain stricter than the state's order level and hence county values may be greater than these values. This is relevant both for simple state-wide orders and for tiered orders. |
| PH_Orders_linklist | Contains url for each of the public health orders included in the dataset, by county and date of order. |

**Appendix Table 2.**
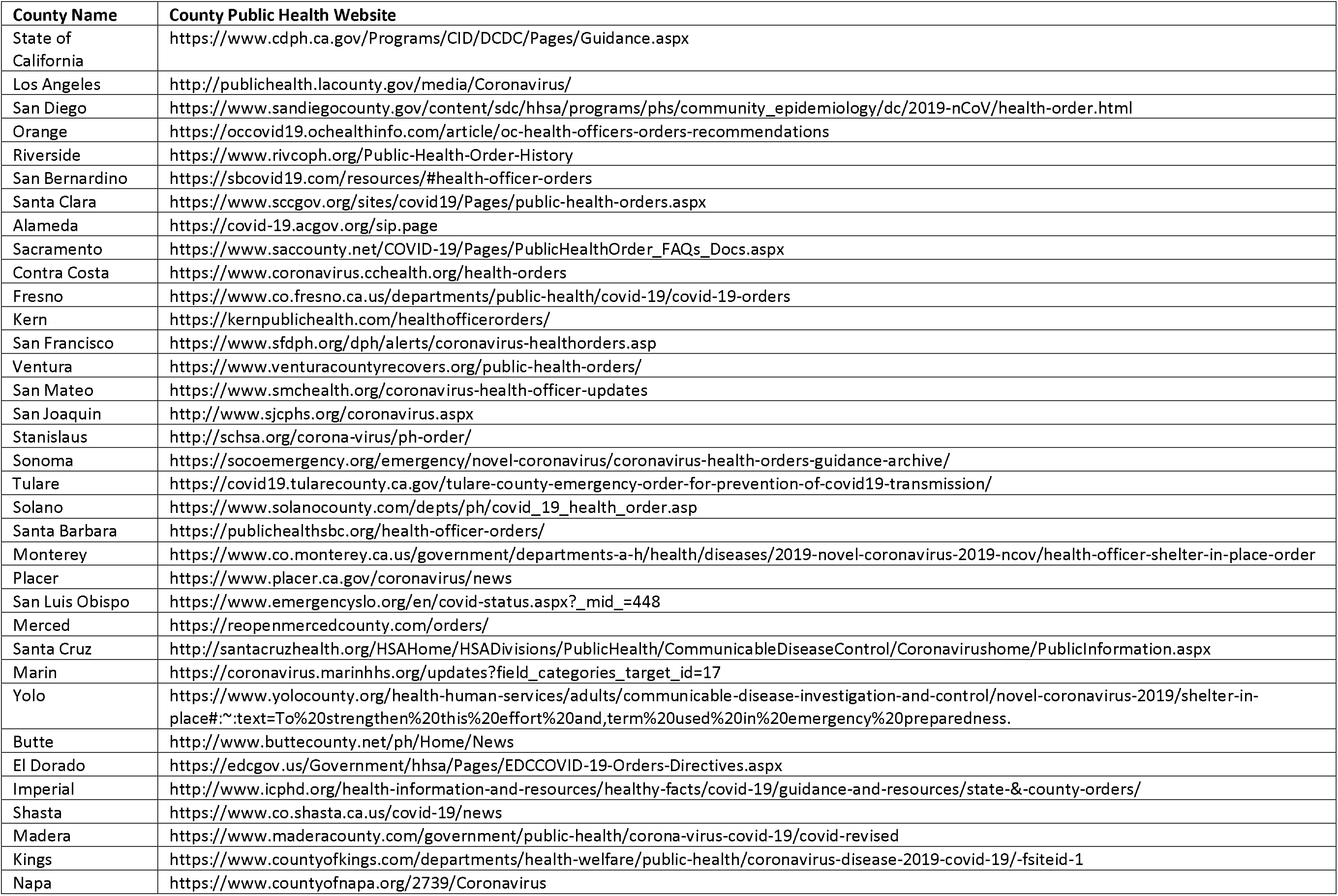
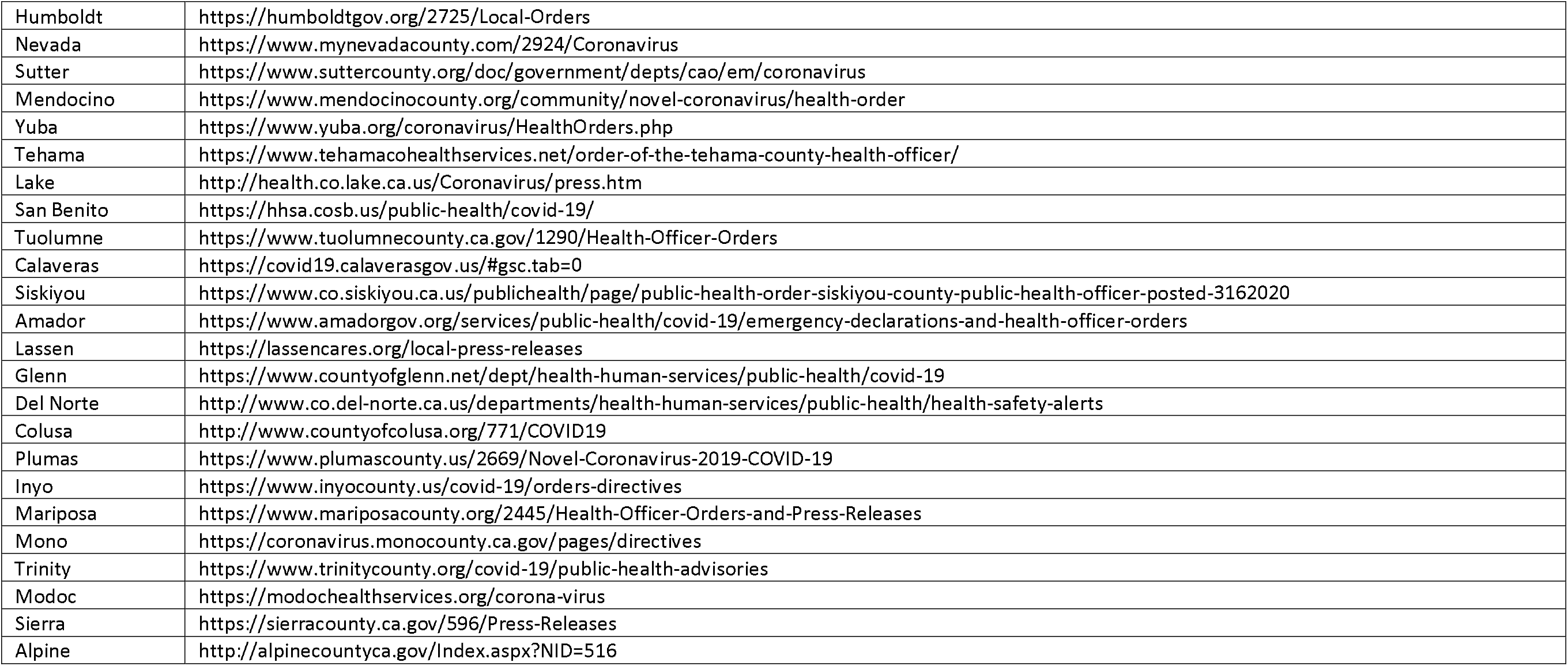
Websites accessed for searches of public health orders (main governmental sites)

<u>Appendix Table 3. Additional websites accessed for county and state public health orders</u>

The list of each public health order accessed and its URL link are now provided as a CSV file with the public data releases. For more information on the structure of the file, see Appendix Table 1.

**Appendix Table 4:**
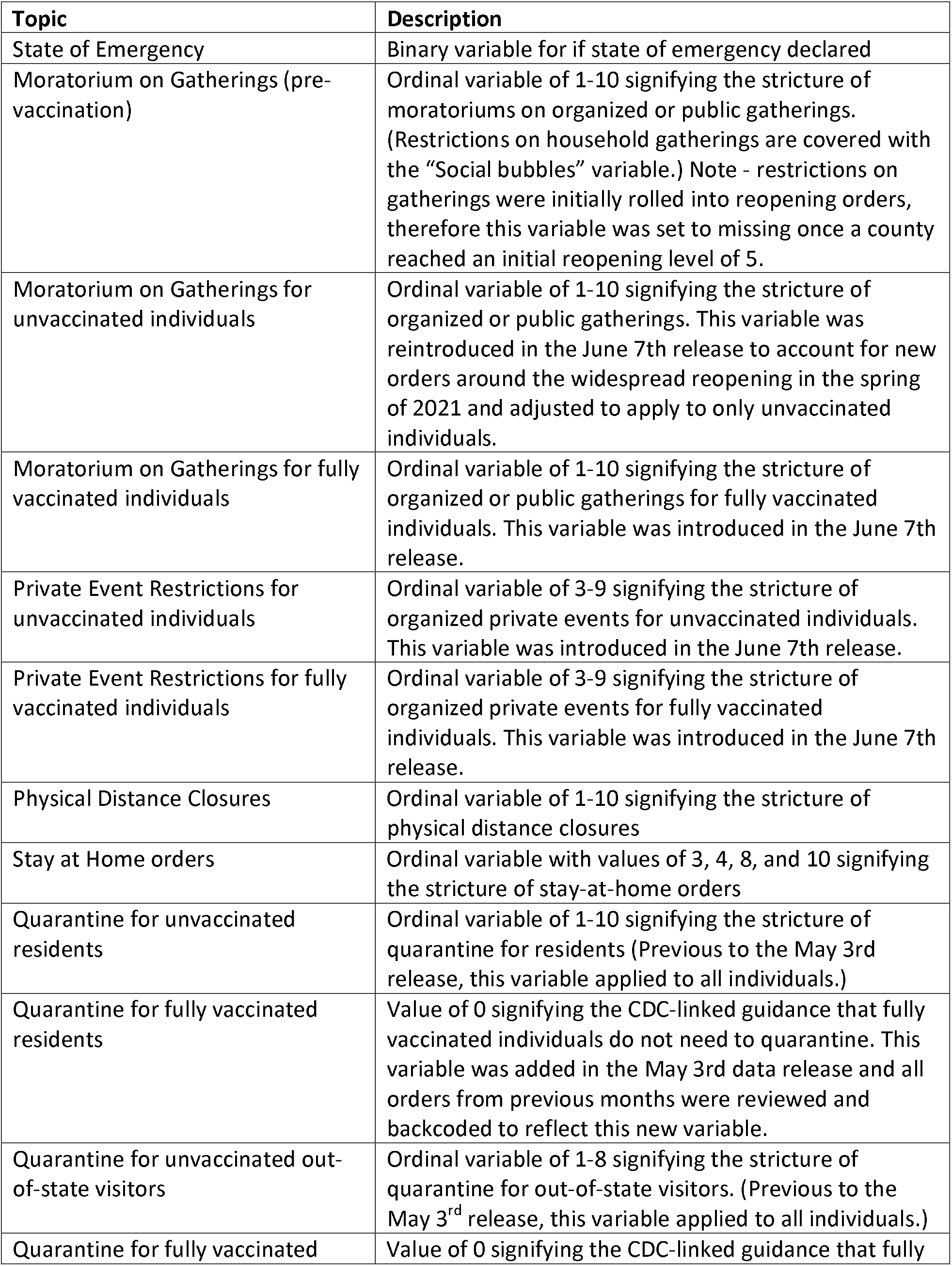
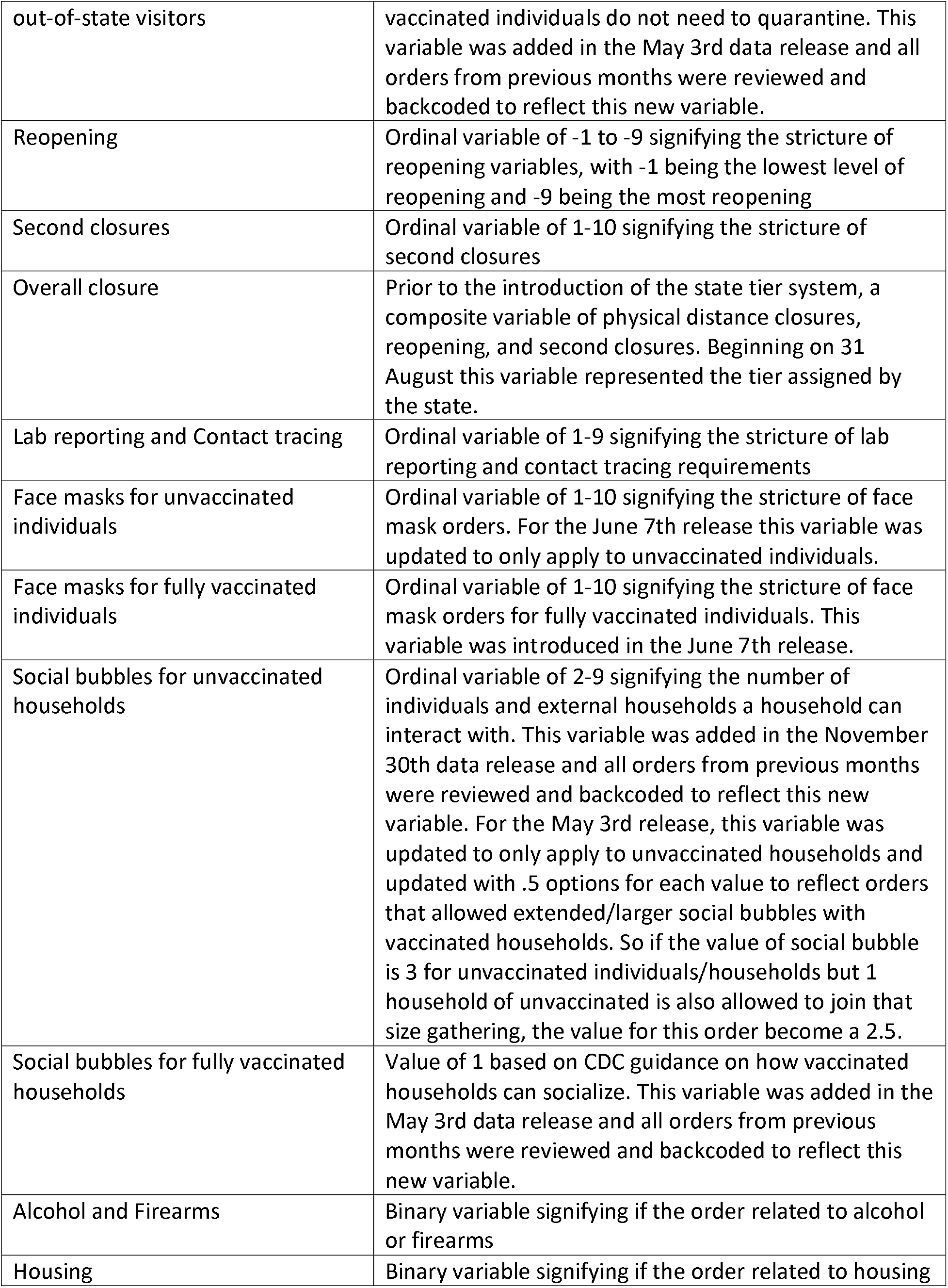
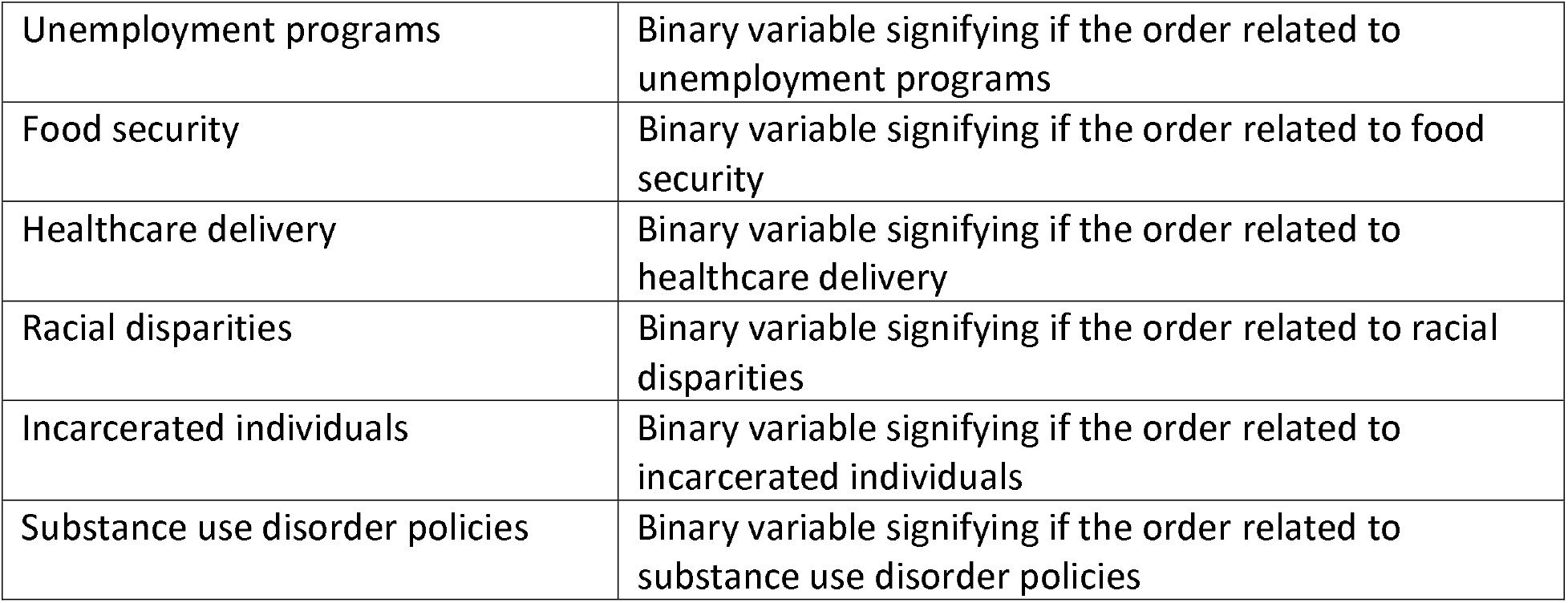
List of topics covered with definition

**Appendix Table 5.**
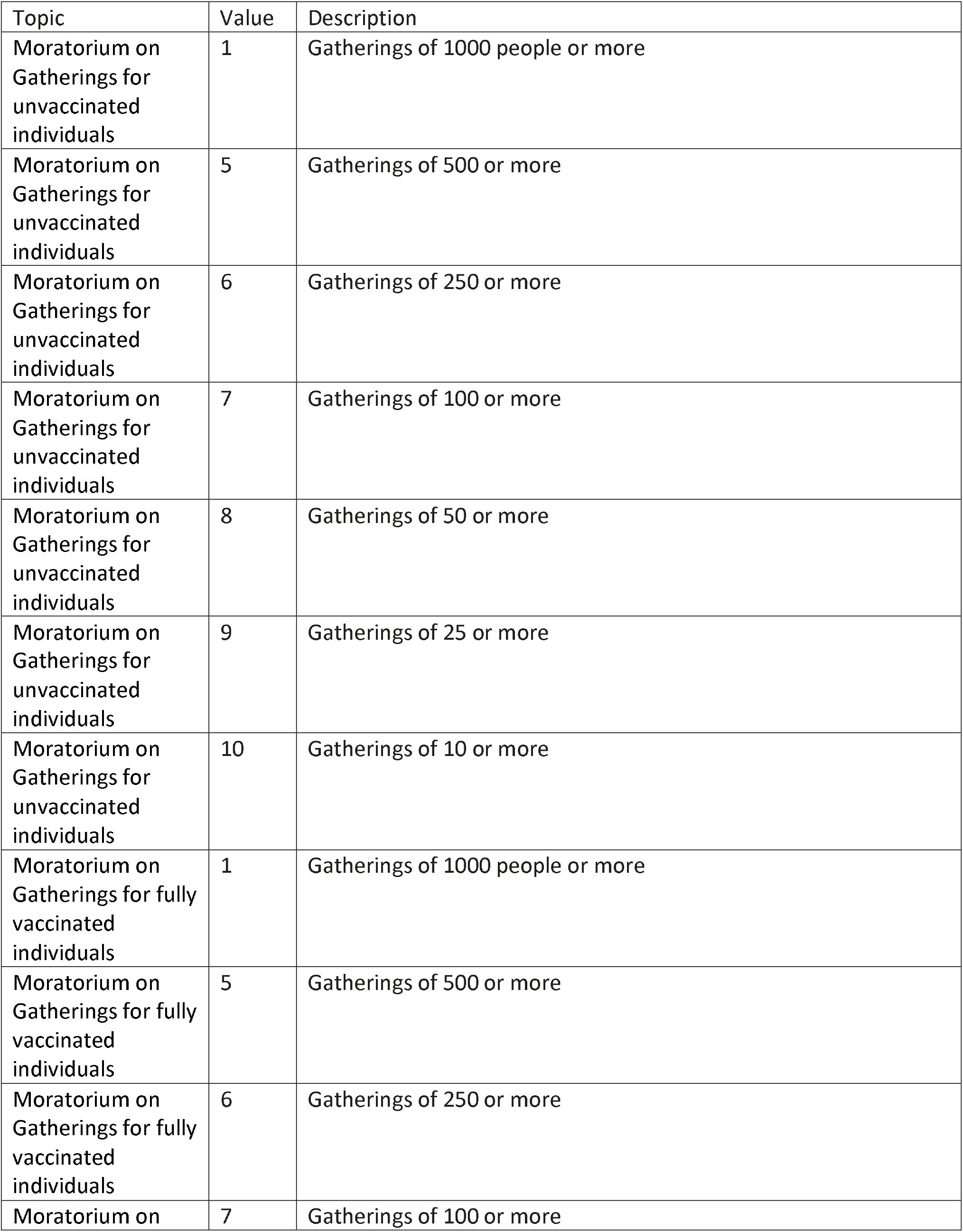
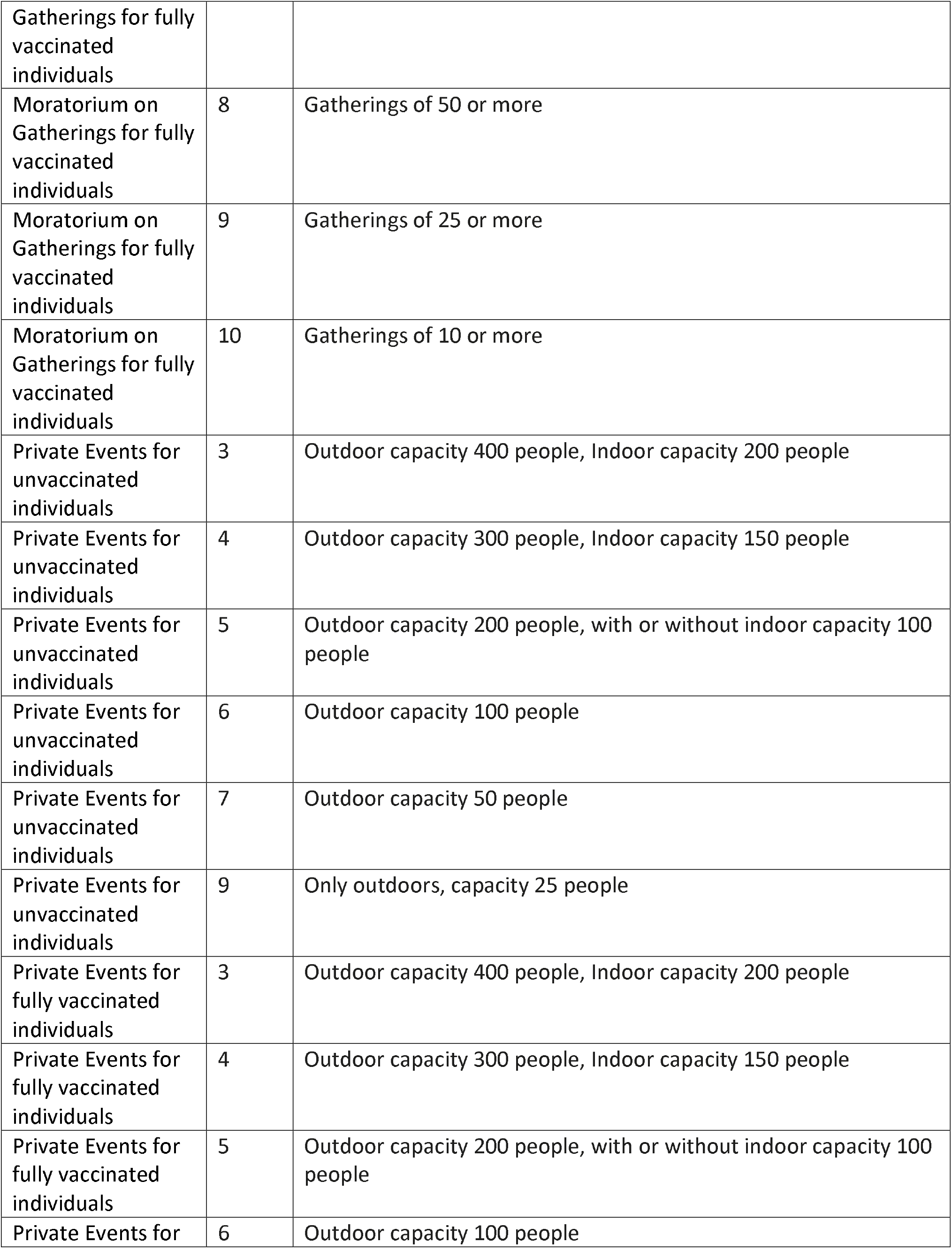
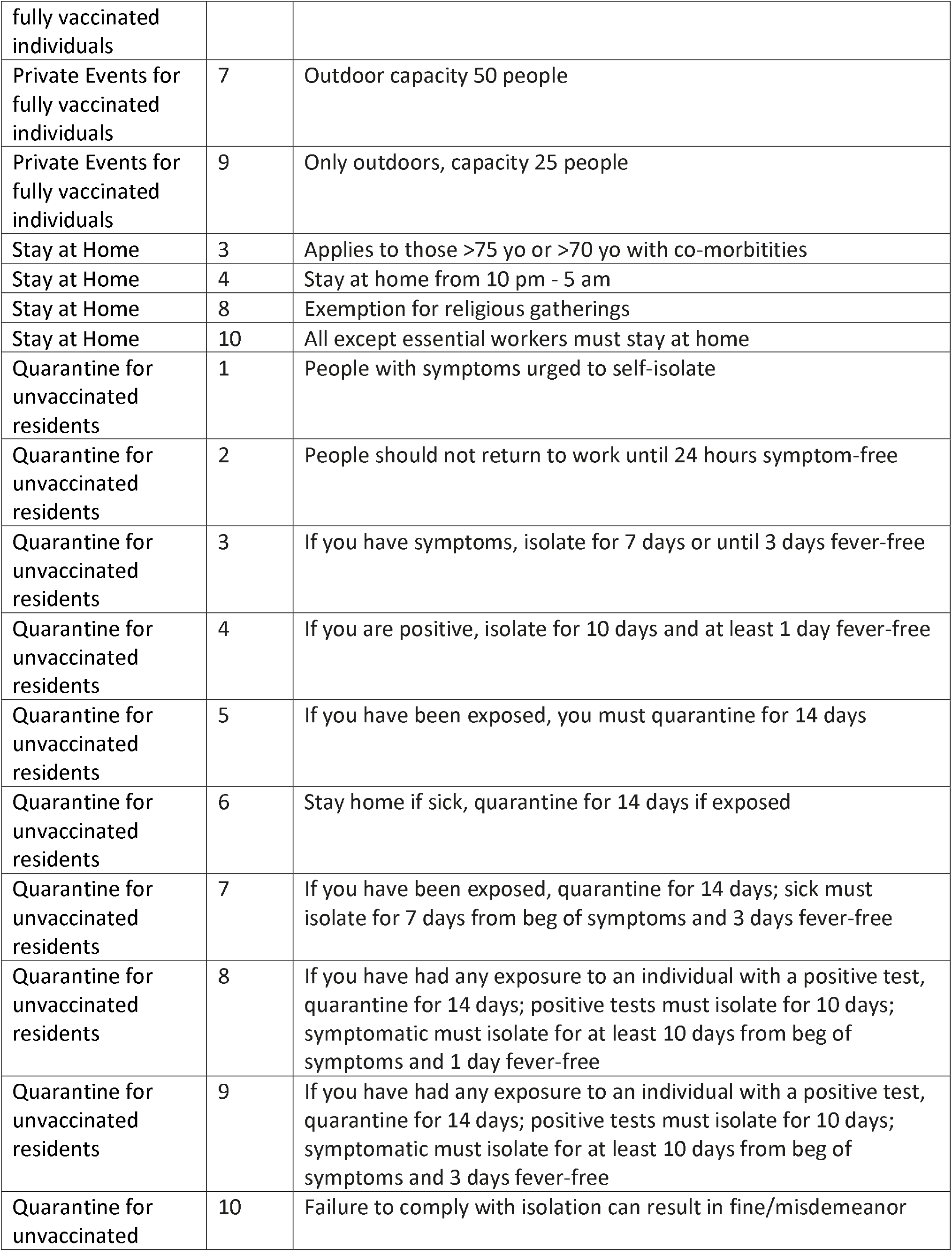
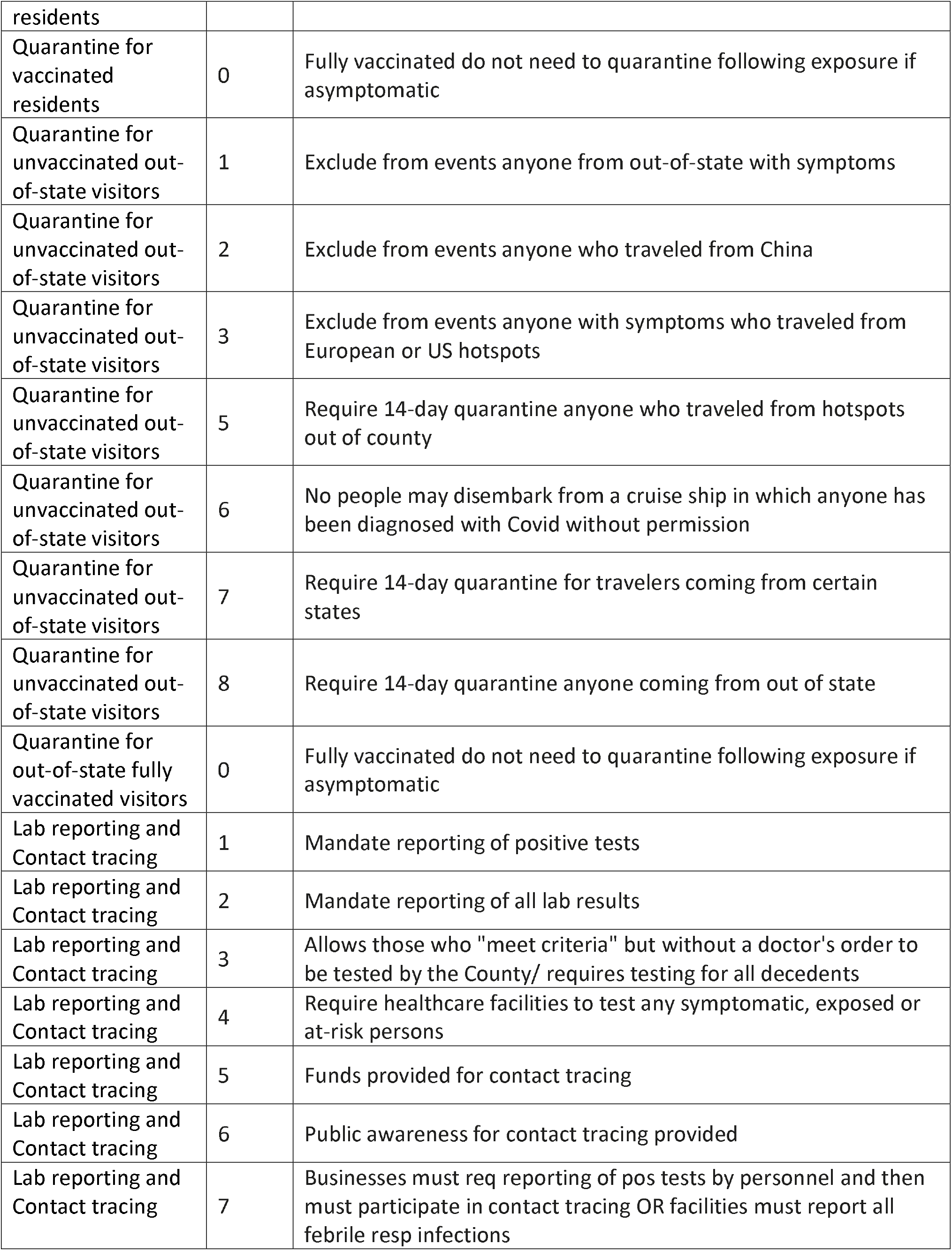
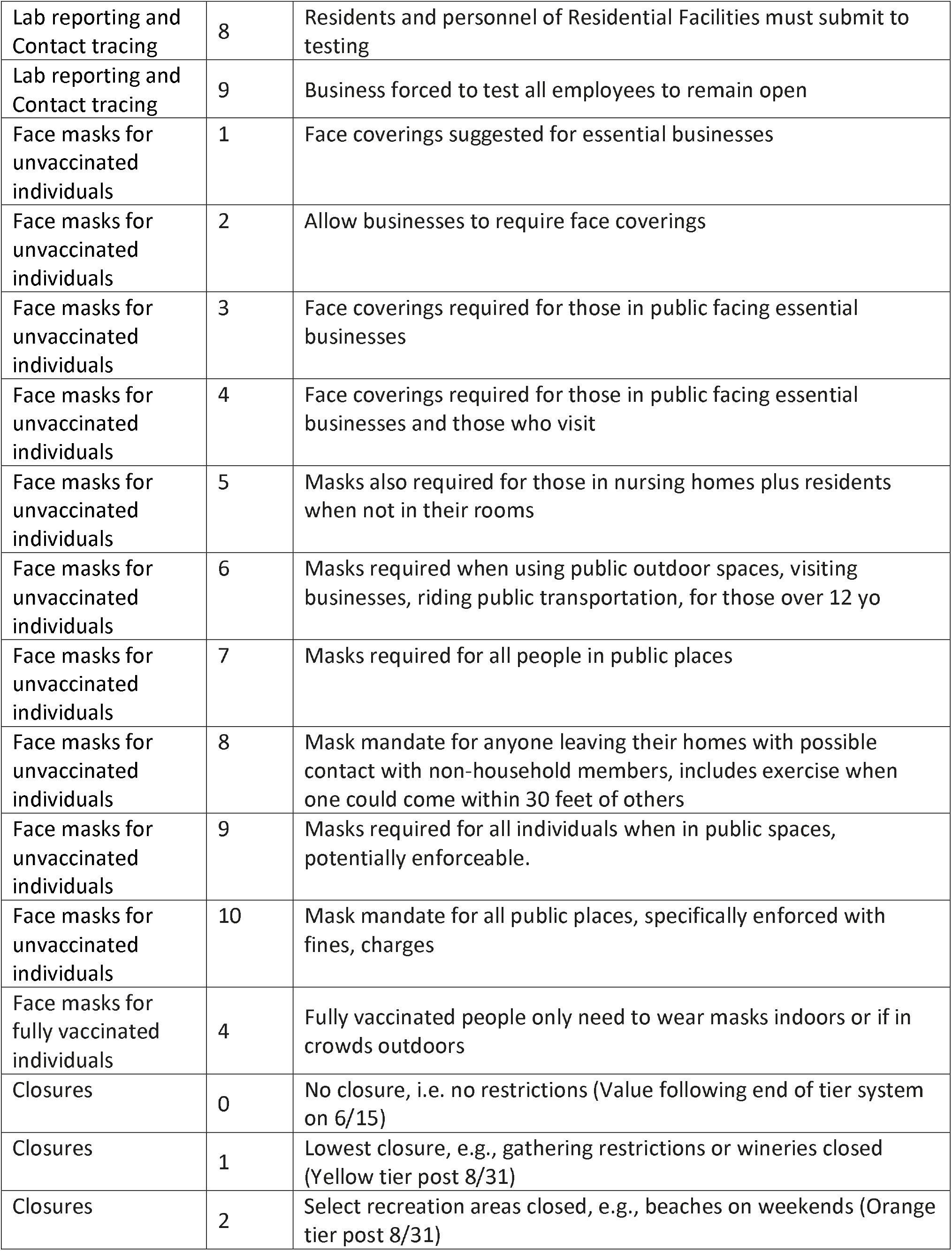
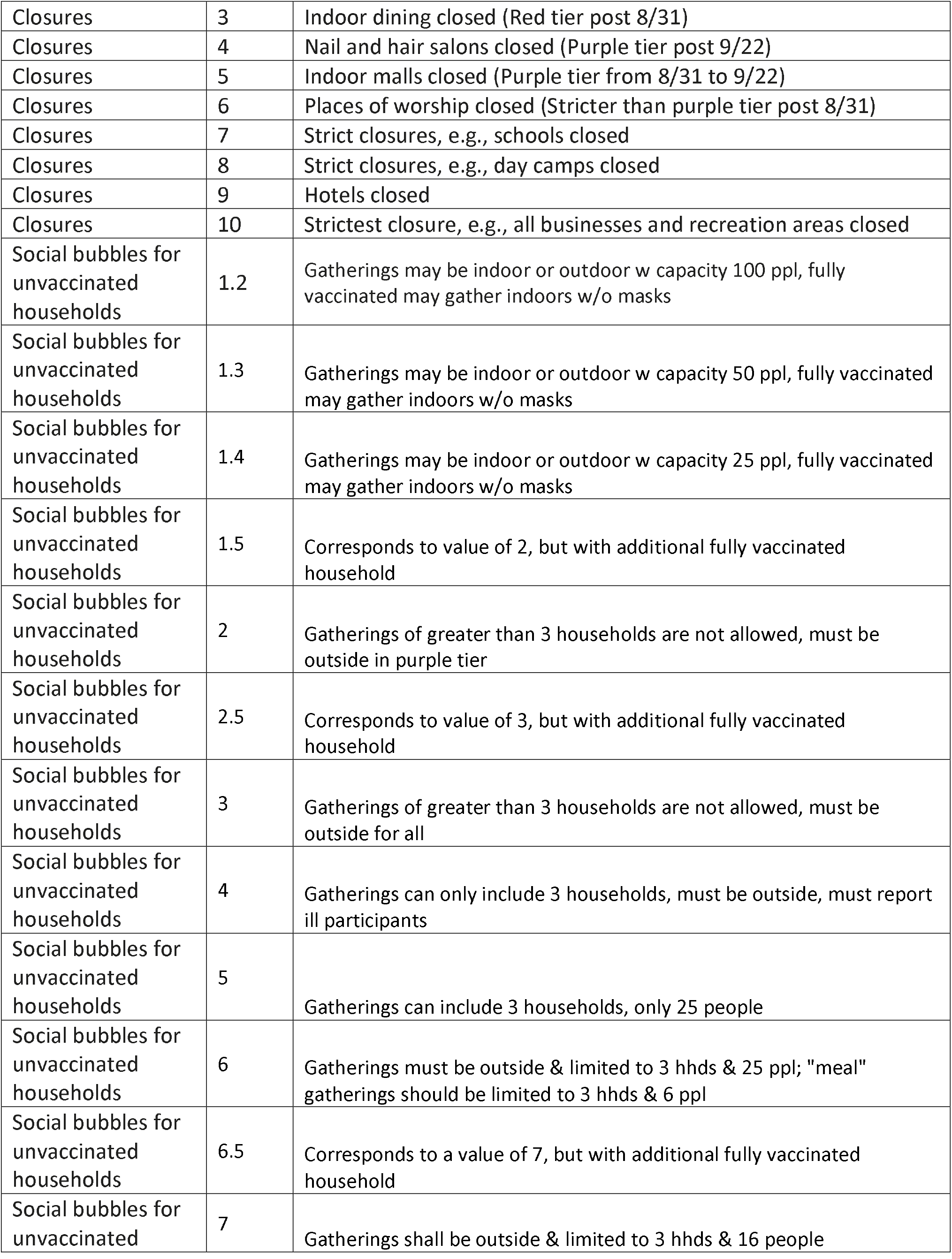
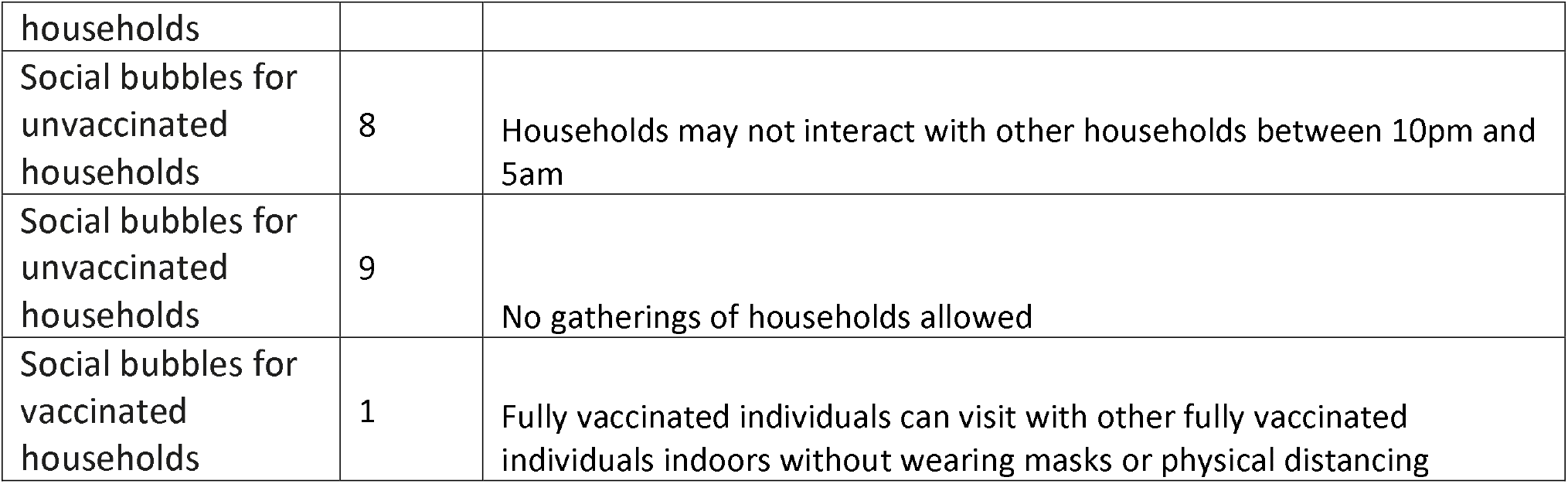
Descriptions of value definitions for each ordinal public health order category

## Footnotes

1 Expiry dates were mainly used by counties in the beginning of the COVID-19 epidemic and were typically overwritten by additional orders before their expiration.

2 The topic of social bubbles, i.e., the number of individuals that your household can interact with, was introduced for the November 30^th^ data release based on new orders being issued on this topic.

3 The topics of quarantine for fully vaccinated residents, quarantine for fully vaccinated visitors, and social bubbles for vaccinated households were introduced for the May 3^rd^ data release based on new orders being issued on these topics. The original topics changed to quarantine instructions for unvaccinated individuals and households. The values for social bubbles for unvaccinated households were expanded to reflect guidance that allowed an additional fully vaccinated household to join social bubbles - more details on the values can be found in Appendix Table 5.

4 The topics of private events for unvaccinated and for fully vaccinated individuals were introduced with the June 7^th^ data release based on new orders being issued on these topics. Additionally, the original topic of gatherings became gatherings for unvaccinated individuals and a new topic of gatherings for fully vaccinated individuals was introduced with the June 7^th^ data release.

## Notes

### Competing Interest Statement

The authors have declared no competing interest.

### Funding Statement

This work was supported in part by contracts with the California Department of Technology and the California Department of Public Health [agreement number 19-13050] and by the Stanford School of Medicine COVID-19 Emergency Response Fund established with generous gifts from donors. Additionally, this work was supported in part by the National Institute On Drug Abuse of the National Institutes of Health [award number R37DA015612]. The content is solely the responsibility of the authors and does not necessarily represent the official views of the National Institutes of Health or of the State of California.

### Author Declarations

As this study uses only publicly available non-individual-level data, it was exempt from IRB review

### Summary of Updates

We have extended our review, categorization, and coding of state- and county-level public health orders through June 30, 2021 and updated the manuscript and appendices accordingly.

